# Timing and Predictors of Loss of Infectivity among Healthcare Workers with Primary and Recurrent COVID-19: a Prospective Observational Cohort Study

**DOI:** 10.1101/2023.06.16.23291449

**Authors:** Stefka Dzieciolowska, Hugues Charest, Tonya Roy, Judith Fafard, Sara Carazo, Ines Levade, Jean Longtin, Leighanne Parkes, Sylvie Nancy Beaulac, Jasmin Villeneuve, Patrice Savard, Jacques Corbeil, Gaston De Serres, Yves Longtin

**Affiliations:** McGill University Faculty of Medicine, Montreal, Canada; Université de Montréal, Faculté de médecine, Montréal, Canada; Laboratoire de Santé Publique du Québec, Sainte-Anne-de-Bellevue, Canada; Institut National de Santé Publique du Québec, Quebec City, Canada; CHU de Québec – Université Laval, Quebec City, Canada; Centre Hospitalier de l’Université de Montréal (CHUM) and CHUM Research Center, Montréal, Canada; Université Laval, Quebec City, Canada; Lady Davis Research Institute, Montreal, Canada; Jewish General Hospital Sir Mortimer B. Davis, Montreal, Canada

**Keywords:** SARS-CoV-2, COVID-19, contagiousness, infectivity, recurrent infection, viral culture

## Abstract

**Background:** There is a need to understand the duration of infectivity of primary and recurrent COVID-19 and identify predictors of loss of infectivity.

**Methods:** Prospective observational cohort study with serial viral culture, rapid antigen detection test (RADT) and RT-PCR on nasopharyngeal specimens of healthcare workers with COVID-19. The primary outcome was viral culture positivity as indicative of infectivity. Predictors of loss of infectivity were determined using multivariate regression model. The performance of the US CDC criteria (fever resolution, symptom improvement and negative RADT) to predict loss of infectivity was also investigated.

**Results:** 121 participants (91 female [79.3%]; average age, 40 years) were enrolled. Most (n=107, 88.4%) had received ≥3 SARS-CoV-2 vaccine doses, and 20 (16.5%) had COVID-19 previously. Viral culture positivity decreased from 71.9% (87/121) on day 5 of infection to 18.2% (22/121) on day 10. Participants with recurrent COVID-19 had a lower likelihood of infectivity than those with primary COVID-19 at each follow-up (day 5 OR, 0.14; p<0.001]; day 7 OR, 0.04; p=0.003]) and were all non-infective by day 10 (p=0.02). Independent predictors of infectivity included prior COVID-19 (adjusted OR [aOR] on day 5, 0.005; p=0.003), a RT-PCR Ct value <23 (aOR on day 5, 22.75; p<0.001), but not symptom improvement or RADT result.

The CDC criteria would identify 36% (24/67) of all non-infectious individuals on Day 7. However, 17% (5/29) of those meeting all the criteria had a positive viral culture.

**Conclusions:** Infectivity of recurrent COVID-19 is shorter than primary infections. Loss of infectivity algorithms could be optimized.

## Introduction

Coronavirus disease 2019 (COVID-19) is caused by the severe acute respiratory syndrome coronavirus 2 (SARS-CoV-2).^1^ The current evidence regarding duration of infectivity rely on viral culture to detect shedding of replication-competent virus (also called viable or infectious virus). These studies suggest that immunocompetent individuals with non-severe COVID-19 can remain infective for up to 10 days.^2–6^

While infective, healthcare workers (HCWs) with COVID-19 must refrain from working to prevent nosocomial transmission.^7, 8^ However, the timing of their return to work is complicated by the interindividual variation in the durations of infectivity. Approximately a fifth of individuals may be infective for as little as 5 days, while approximately a quarter can remain infective for 10 days or more.^9^ Determinants of loss of infectivity are largely unknown, but could be useful to optimize the return-to-work of infected HCWs. To limit absenteeism,^10^ the US Centers for Disease Control and Prevention (CDC) and European CDC have provided guidance to allow earlier return to work of eligible HCWs.^7, 8^ These algorithms use readily available information such as symptom improvement and the result of rapid antigen detection tests (RADT) to predict loss of infectivity.^7, 8^ However, whether these criteria can reliably distinguish infective and non-infective individuals remain unclear.

Furthermore, many studies that investigated duration of infectivity were conducted early in the pandemic when individuals were infected for the first time, and often were unvaccinated. Few studies have investigated duration of infectivity of recurrent COVID-19.^9^ Hence, we sought to evaluate the duration of infectivity of HCWs infected with primary and recurrent COVID-19, and identify predictors of infectivity using viral culture as a marker of infectivity.

## Methods

### Study population

We conducted a prospective observational cohort study at the CIUSSS Centre-Ouest-de-Montréal, a large healthcare organization employing 12,000 HCWs across 20 institutions. Participants were identified through the Occupational Health Service. Inclusion criteria included (1) SARS-CoV-2 infection confirmed by reverse transcription polymerase chain reaction (RT-PCR) from a nasopharyngeal specimen, and (2) symptom onset <72h prior to enrolment. Exclusion criteria included asymptomatic infections; moderate-to-severe COVID-19 (WHO Ordinal Scale for Clinical Improvement >=3)^11^; contraindication to nasopharyngeal sampling; and use of COVID-19-specific therapies (e.g. antivirals or monoclonal antibodies). Participants were followed on the 5^th^, 7^th^ and 10^th^ day of their infection (with the day of onset of symptoms defined as day 1).

The study follows the Strengthening the Reporting of Observational Studies in Epidemiology (STROBE) guideline,^12^ and was approved by local research ethics committees. Written informed consent was obtained from participants.

### Data collection

Clinical data included sociodemographic information, medical history (including prior COVID-19 infection), COVID-19 vaccination status (including number of doses and manufacturer), and symptomatology. We also assessed the use of antipyretics (acetaminophen and non-steroidal anti-inflammatory drugs) among afebrile participants as their use can mask fever. Participants reported this information online (LimeSurvey GmbH, Hamburg, Germany).

### Outcome definitions

SARS-CoV-2 infectivity was defined as evidence of cytopathic effect (CPE) on microscopy of viral culture from a nasopharyngeal specimen, with etiology of the CPE being confirmed as SARS-CoV-2 by RT-PCR on the culture supernatant demonstrating at least 3 cycle threshold (Ct) values lower than the RT-PCR on the original sample.^13^ Duration of infectivity was defined as the number of days between the onset of symptoms and the last positive culture.

### Laboratory methods

Nasopharyngeal samples using a flocked swab (FLOQSwabs, Copan Italia) were placed in 3ml universal viral transport media (UTM, Copan Italia) and kept at -80°C. Primary samples and culture supernatants were processed with an in-house RT-PCR targeting the SARS-CoV-2 N gene as previously described.^14^ Forward, reverse and probe sequences were as follows: AACCAGAATGGAGAACGCAGTG, CGGTGAACCAAGACGCAGTATTAT and CGATCAAAACAACGTCGGCCCCAAGGTTTAC .^14^

Viral cultures were performed on Vero E6 cells as previously described.^14^ This cell line is commonly used to cultivate SARS-CoV-2 and has a median tissue culture infectious dose (TCID_50_) ranging between 2.0E+04 to 6.3E+06.^15^ A 0.1 ml aliquot of specimen was used as an inoculum. Cultures were kept for 15 days.

All initial samples were sequenced to determine SARS-CoV-2 lineage using the Illumina technology. Data analysis was performed using the GenPipes Covseq pipeline^16^ and variant identification was performed with Pangolin program (see appendix for details)^17^.

Lateral-flow RADT were provided for self-administration (COVID-19 Antigen Rapid Test, BTNX, Hannover, Germany). Participants performed the test on a self-sampled midturbinate swab specimen by following the manufacturer’s instructions and interpreted the test result as positive, negative, or uncertain.^18^

### Sample size estimate

Based on studies indicating that 25% of individuals remain infective on the 7^th^ day of their infection,^19^ we estimated that recruiting 120 participants would provide a precision of +/-7% at the 95% confidence interval (CI).

### Statistical analyses

Descriptive statistics reported discrete variables as numbers and proportions, and continuous variables as mean ± standard deviation (SD) or median and interquartile ranges (IQR). The primary outcome was the proportion of HCWs with evidence of infectivity on the 5^th^, 7^th^ and 10^th^ day of their infection.

To investigate the capacity of the RT-PCR Ct value (an indicator of viral load that is inversely proportional to the quantity of nucleic acid in a sample) and RADT to predict infectivity, the Ct values of RT-PCR of samples with positive vs negative viral culture was depicted in the form of boxplots with overlaid jitter plot.

To investigate factors associated with persistent infectivity, odds ratios (OR) and 95% confidence intervals (CI) were estimated using bivariate and multivariate logistic regression at each follow-up visit. Multivariate models included clinical characteristics (symptom severity, symptom resolution, and fever) and results laboratory assays (RADT and RT-PCR) collected at each follow-up, as well as baseline individual (age, sex and immunological status) and viral (SARS-CoV-2 lineage) information. These variables were pre-defined as potential predictors of duration of infectivity according to literature and current practices.^7, 8^ Categories were grouped when necessary for model convergence. Variables perfectly predicting the presence of infectivity could not be included in the corresponding multivariate model. Immunologic status was categorized according to vaccination and prior infection as follows: recent vaccination (last dose received <6 months ago) without prior infection; non-recent vaccination (last dose received ≥6 months ago) without prior infection; and hybrid immunity (vaccination at any time and prior infection).

### Performance of return-to-work algorithms

We estimated the capacity of the CDC algorithm to discriminate infective and non-infective HCWs on day 7 of their infection.^7^ We also quantified the probability of an infectious HCW returning to work, and estimated the impact of these criteria to limit absenteeism. Finally, we explored the performance of alternative algorithms to predict loss of infectivity that were derived from variables identified in the current study. These evaluations assumed that in the absence of return-to-work criteria, HCWs would return to work 10 days after the onset of their symptoms.

All analyses were performed in SAS version 9.4. All tests were 2-tailed and a p-value < 0.05 was considered statistically significant. Adjustment for multiple comparisons were not applied in this exploratory study.^20, 21^

### Role of Funding source

The study was sponsored by the Ministère de la Santé et des Services Sociaux du Québec. The sponsor had no role in the design, conduct and reporting of the study.

## Results

Between Feb 20^th^ 2022 and March 6^th^, 2023^th^, 237 patients were offered to participate, and 121 (51.1%) were included in the analyses (Figure 1). Overall, 714 specimens (360 nasopharyngeal and 354 mid-turbinate swabs) were collected. Baseline characteristics of participants are shown in Table 1; 79.3% (96/121) were female, and the average age was 40.2 (SD, 12.0 years). The infections were due to multiple Omicron lineages including BA.1 (11.6%), BA.2 (60.3%), and BA.5 (8.3%), inclusive of sublineages. Virtually all participants (98.3%) were previously immunized with ≥ 1 dose of SARS-CoV-2 vaccine. Most (84.3%) had received 3 doses, most commonly the Pfizer–BioNTech Comirnaty (89.3% of all received doses). The median elapsed time between the last dose and the current infection was 122 days (IQR, 95-175 days). Twenty (20) participants (16.5%) had a prior COVID-19 episode. All these previous episodes were mild (WHO Grade scale ≤2) and occurred a median of 347.5 days prior to the current episode (IQR, 264 to 454 days).

**FIGURE 1.**
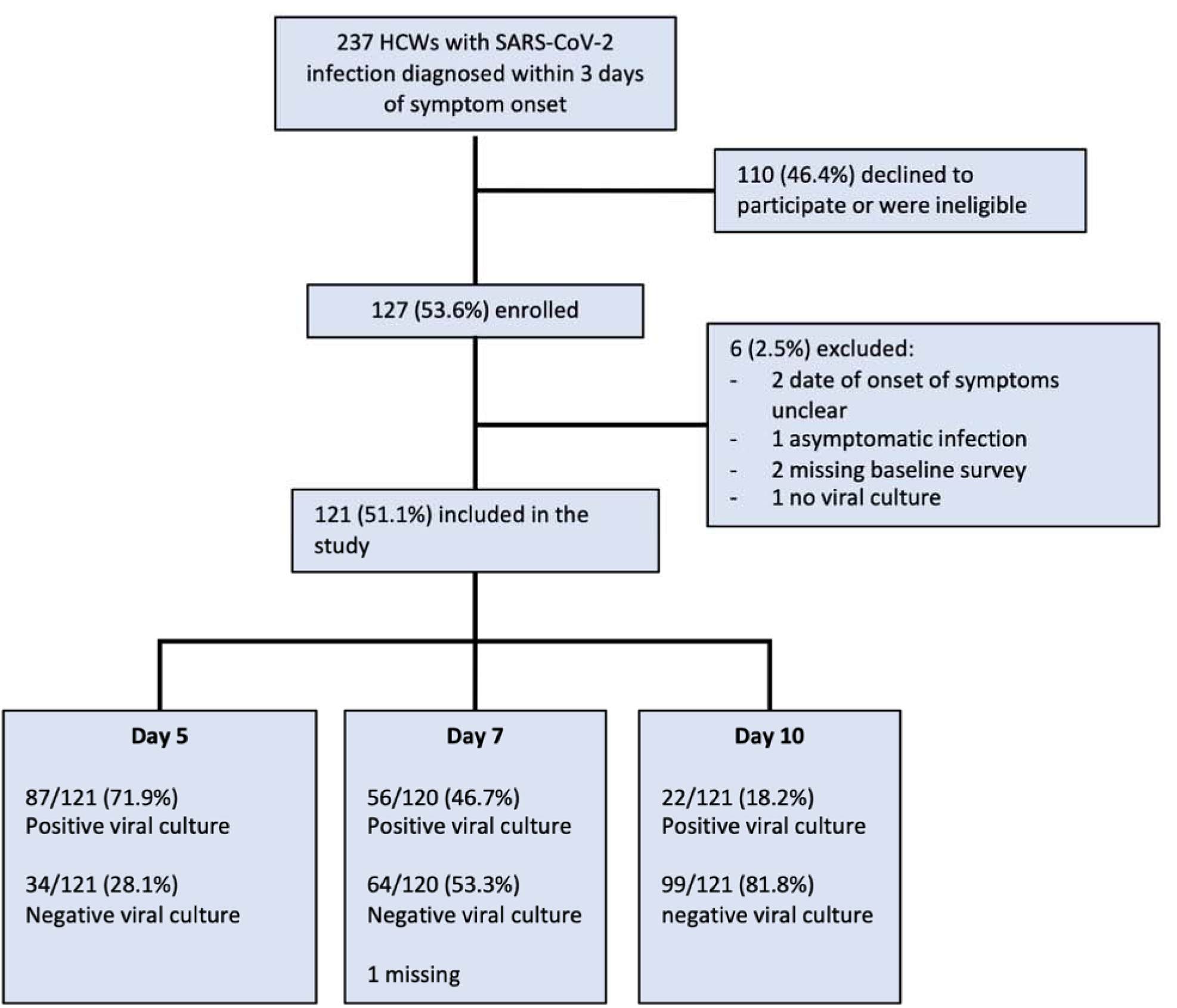
Flow diagram of participant selection into the study and proportion of infective participants at each follow-up visit.

**Table 1.**
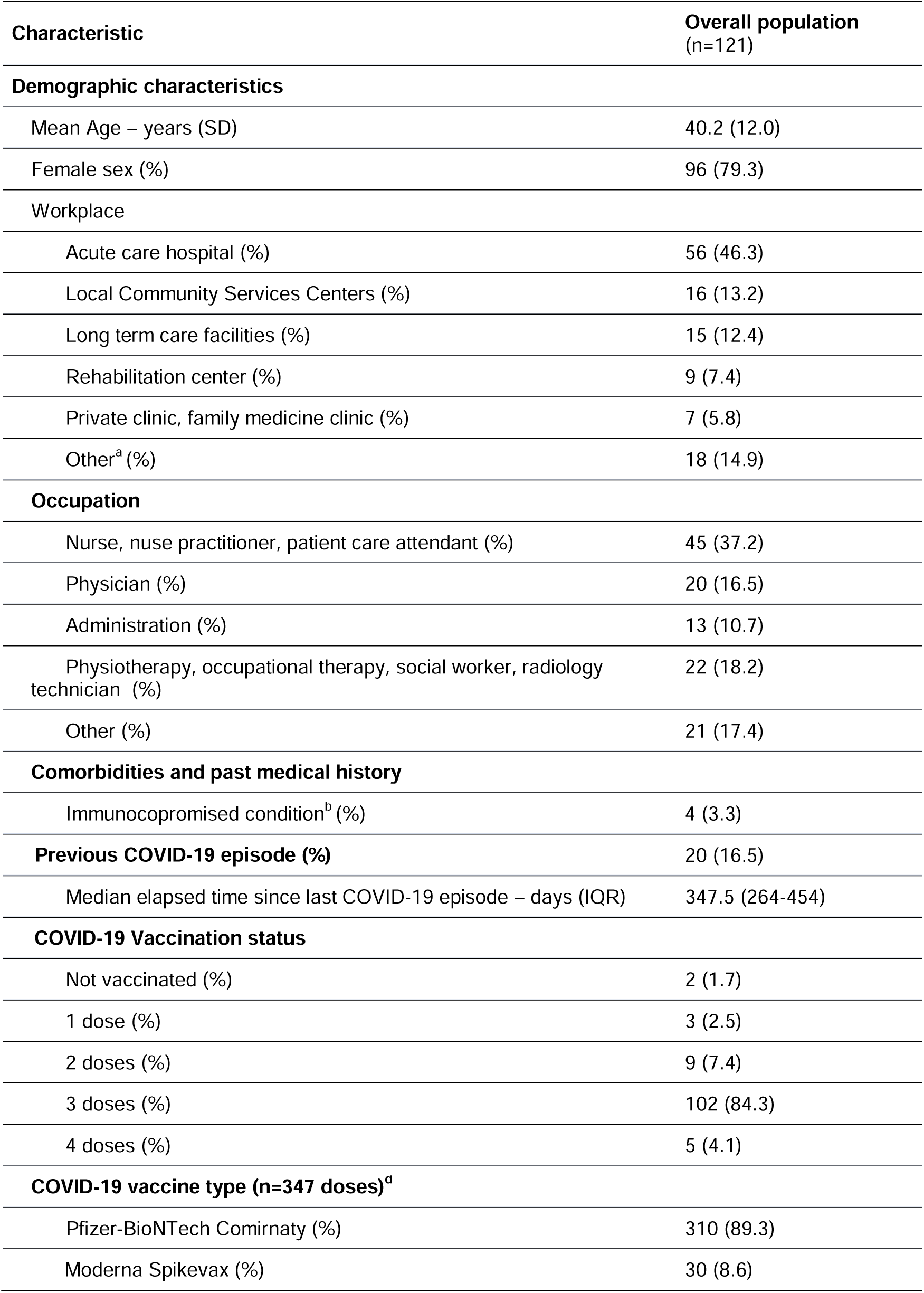
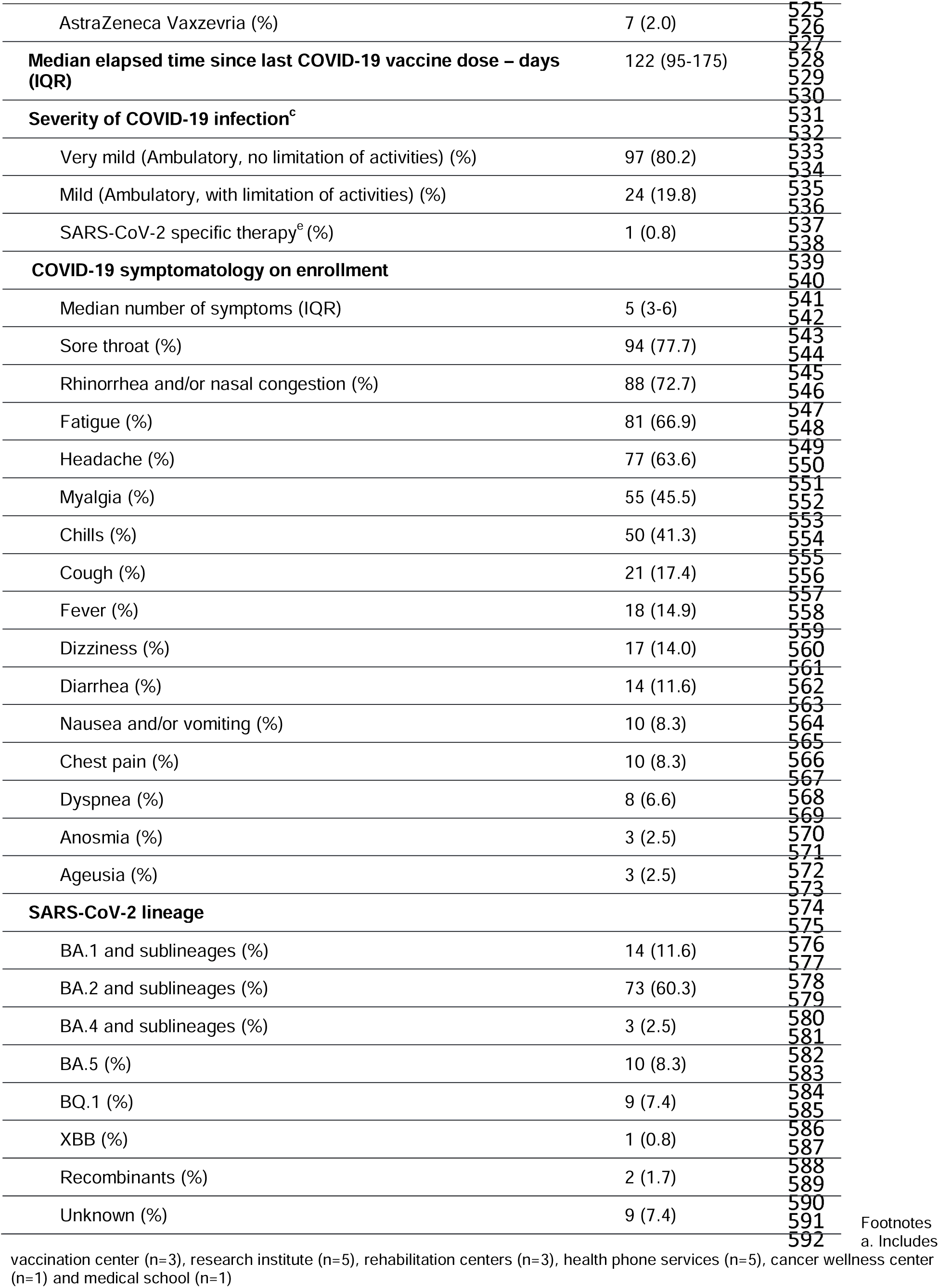
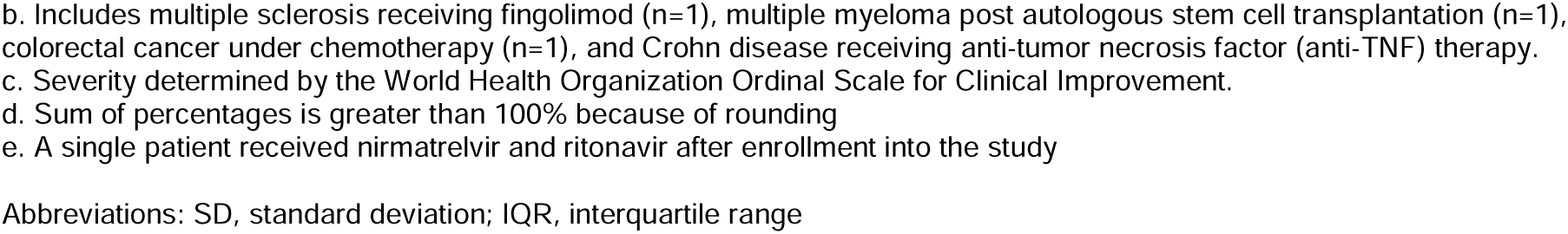
Demographic and clinical characteristics of healthcare workers with COVID-19

### Symptoms of current COVID-19

Upon enrollment, all participants described their infection as either “very mild” or “mild” (WHO Grade scale of 1 or 2). The most common symptoms were sore throat (77.7%), rhinorrhea and/or nasal congestion (72.7%) and fatigue (66.9%). The clinical evolution was favourable. No participant was hospitalized. A single participant received nirmatrelvir-ritonavir after enrollment. The proportion of participants with markedly improved or resolved symptoms increased from 43.8% on day 5 to 84.3% on day 10 (data not shown). Fever was uncommon: only 14.9% were febrile on enrollment. However, antipyretic use was frequent in afebrile individuals. For example, they were used by 50% and 31% of afebrile individuals on day 5 and 7 of their infection, respectively.

### Evolution of infectivity and viral shedding

The proportion of participants with a positive viral culture was 71.9% (87/121; 95% CI, 63.0% to 79.7%) on day 5, 46.7% (56/120; 95% CI, 37.5% to 56.0%) on day 7 and 18.2% (22/121; 95% CI, 11.8% to 26.2%) on day 10, respectively (Figure 1). The proportion of participants with a positive RT-PCR decreased from 93.3% (112/120) on day 5 (median Ct value, 23.4 (IQR, 20.6-27.9)) to 61.2% (74/120) on day 10 (median Ct value, 32.5 (IQR, 28.5 to undetectable)). Similarly, the proportion of RADT tests that were positive decreased progressively from 81.5% (97/119) on day 5, to 34.2% (40/117) on day 10.

### Factors associated with infectivity

In bivariate analysis, multiple variables were associated with a positive viral culture (Table 2). A history of previous COVID-19 was strongly associated with a decreased likelihood of infectivity at each follow-up visit. Only 35% (7/20) of individuals with recurrent COVID-19 were still infective on day 5 compared with 79% of those with a primary episode (OR, 0.14; 95% CI 0.05-0.40; p<0.001). Similarly, only 5% (1/20) of participants with recurrent COVID-19 were still infective on day 7, compared with 55% (55/100) of those with a first episode (OR, 0.04; 95% CI, 0.01-0.33; p=0.003). Finally, the proportion of participants with primary vs recurrent infection that were still infective on day 10 were 22% vs. 0%, respectively (p=0.02 by Fisher’s).

**Table 2.**
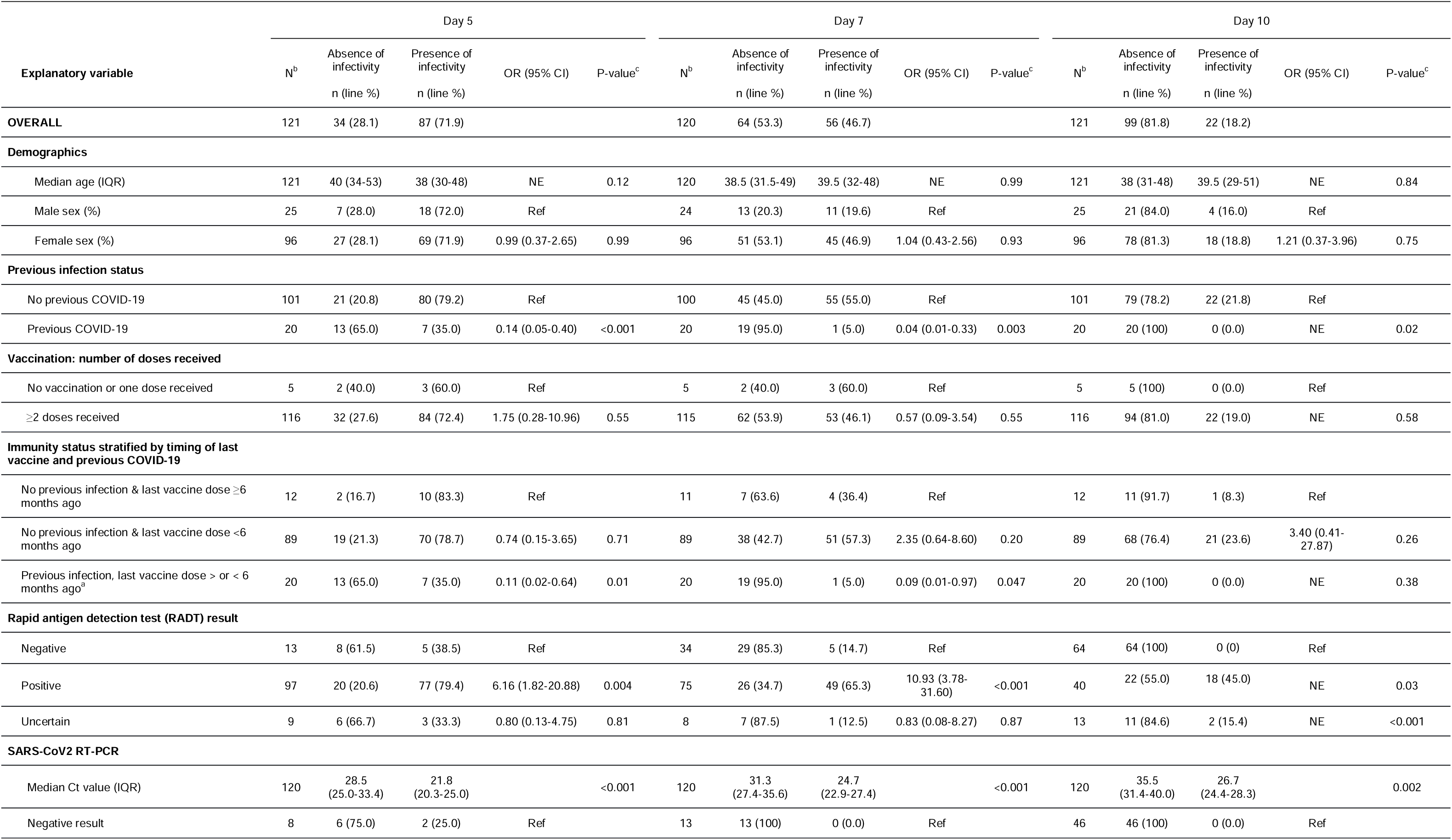
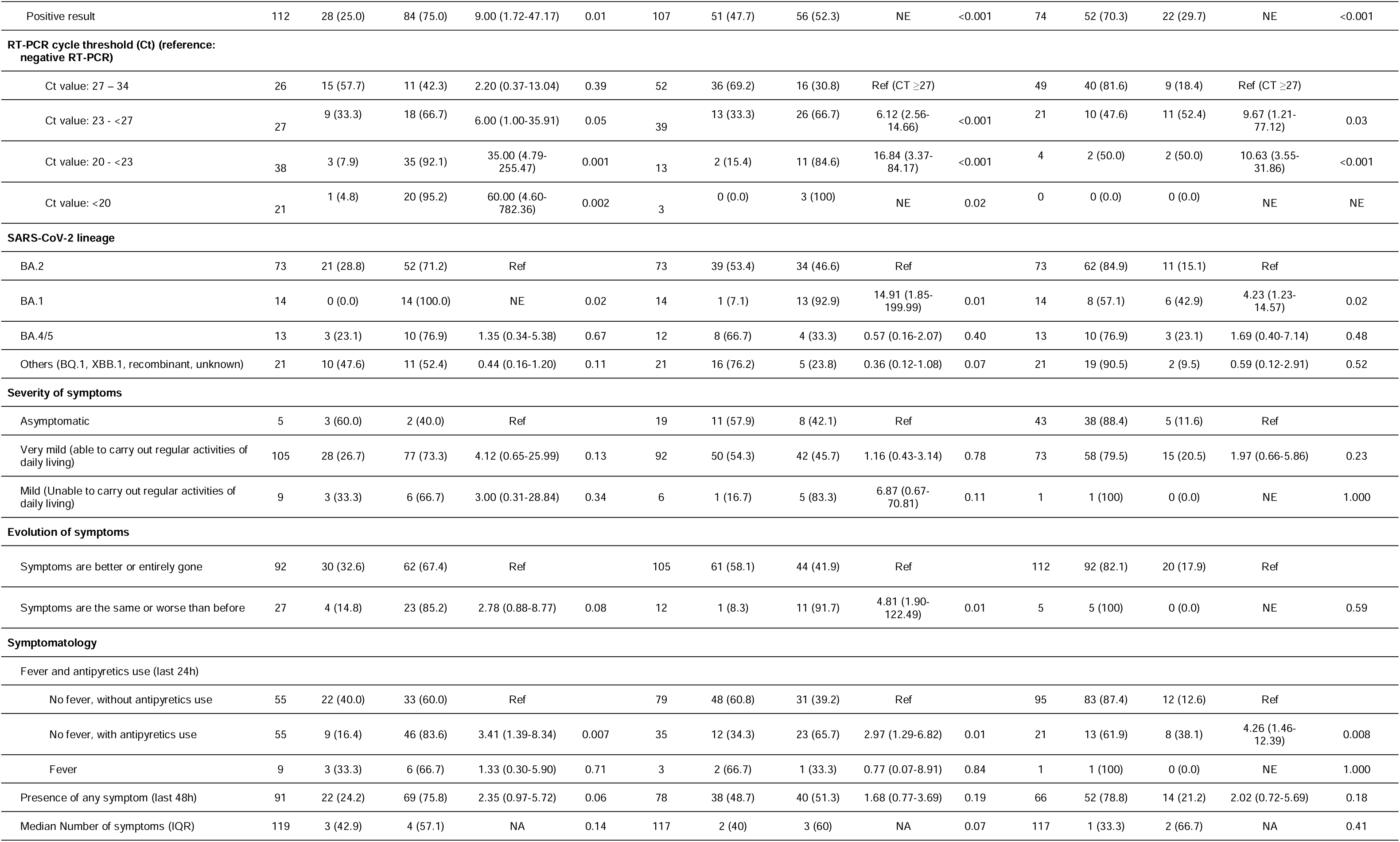
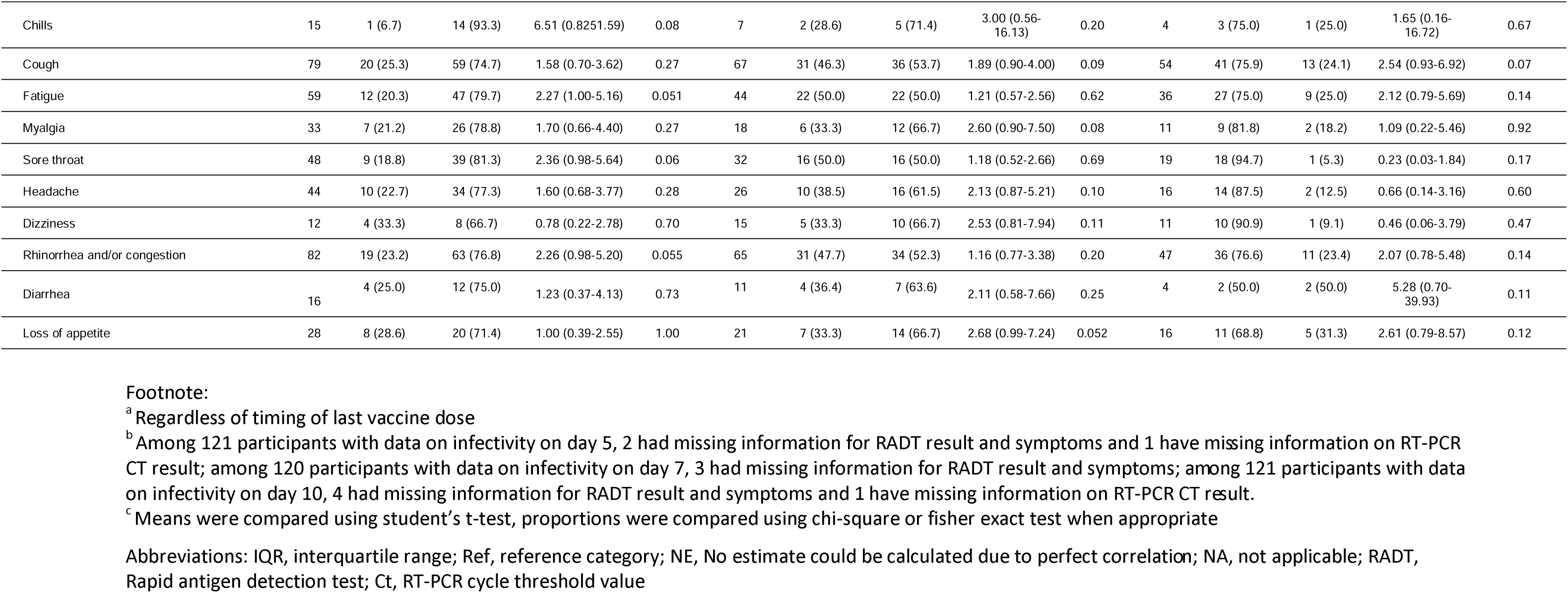
Predictors of infectivity on day 5, 7 and 10 of COVID-19 among healthcare workers (bivariate analyses)

In terms of lineage, the BA.1 lineage was associated with a higher likelihood of infectivity on each follow-up visit than the BA.2 (p ≤0.02), while no difference was detected between BA.2 and BA.4/5.

In terms of clinical features, a lack of symptom improvement was predictive of ongoing infectivity on day 7 (OR, 4.81; p=0.01) but not on day 5 or 10. Also, when compared to afebrile individuals who were not using antipyretics, those who were still using antipyretics were more likely to be infective at each follow up visit (range of OR, 2.97 to 4.26; p≤0.01 for each comparison).

In terms of laboratory assays, a positive RADT result was associated with a higher likelihood of infectivity at day 5 (OR, 6.16; p=0.004) and day 7 (OR, 10.93; p<0.001). A positive RT-PCR was also significantly associated with infectivity at each follow-up, and there was an inverse association between the RT-PCR Ct value and ongoing infectivity (Figure 2). Notably, participants with recurrent COVID-19 differed from those with primary infection in terms of laboratory assays. At each visit, they had significantly higher RT-PCR Ct values (Figure 3) and were significantly more likely to have a negative RADT test result (Table 3).

**FIGURE 2.**
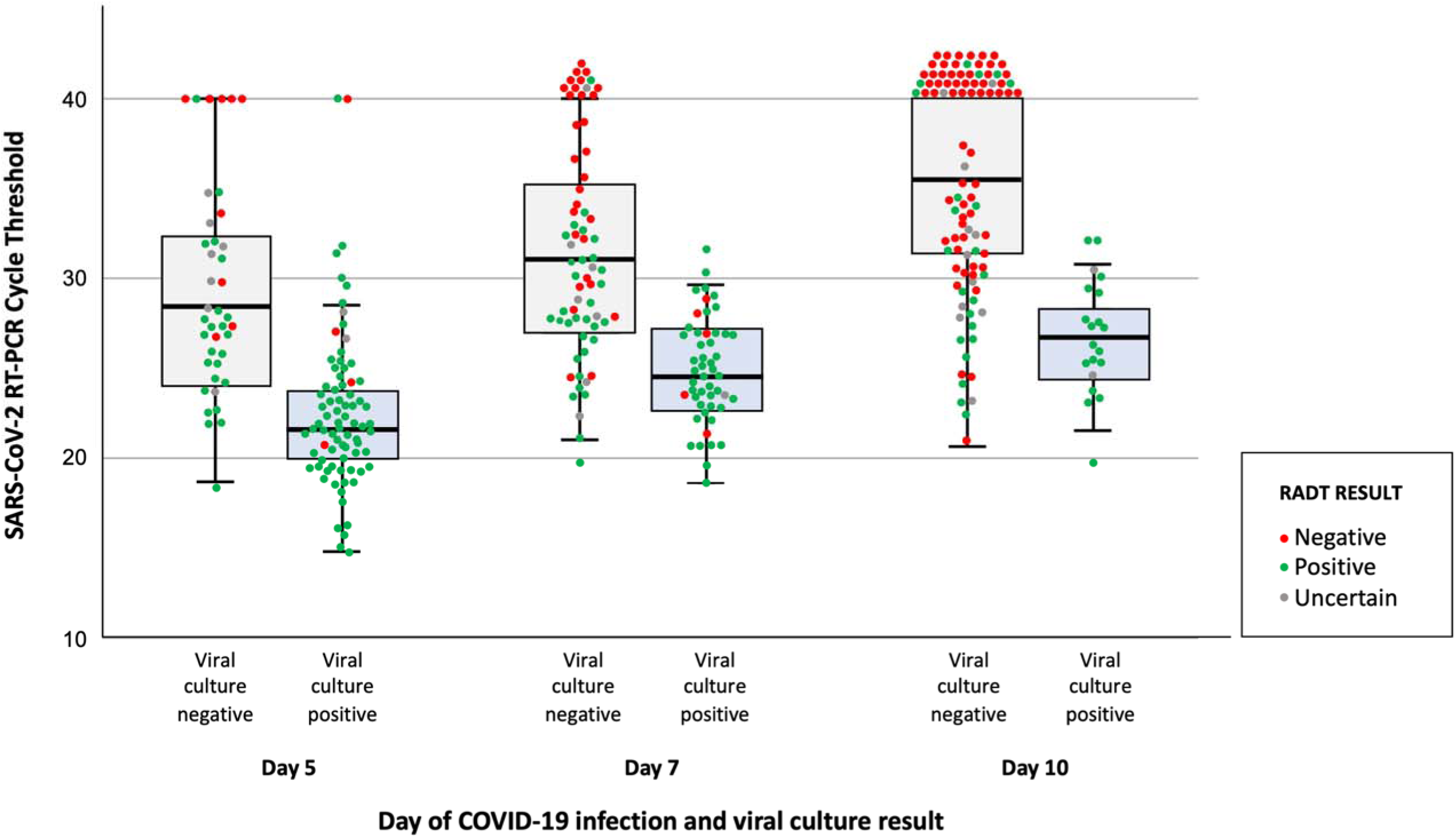
Box plot with overlaid jitter plot comparing SARS-CoV-2 RT-PCR Cycle threshold, rapid antigen diagnostic test (RADT) result and viral culture positivity at day 5, 7 and 10 of COVID-19 among 121 healthcare workers. Footnote: The horizontal line in each box indicates the median, whereas the top and bottom of the boxes represent the 75th and 25th percentile, respectively. Error bars represent 95% confidence intervals. Negative RT-PCR results were attributed a Ct-value of 40 to facilitate data visualization.

**FIGURE 3.**
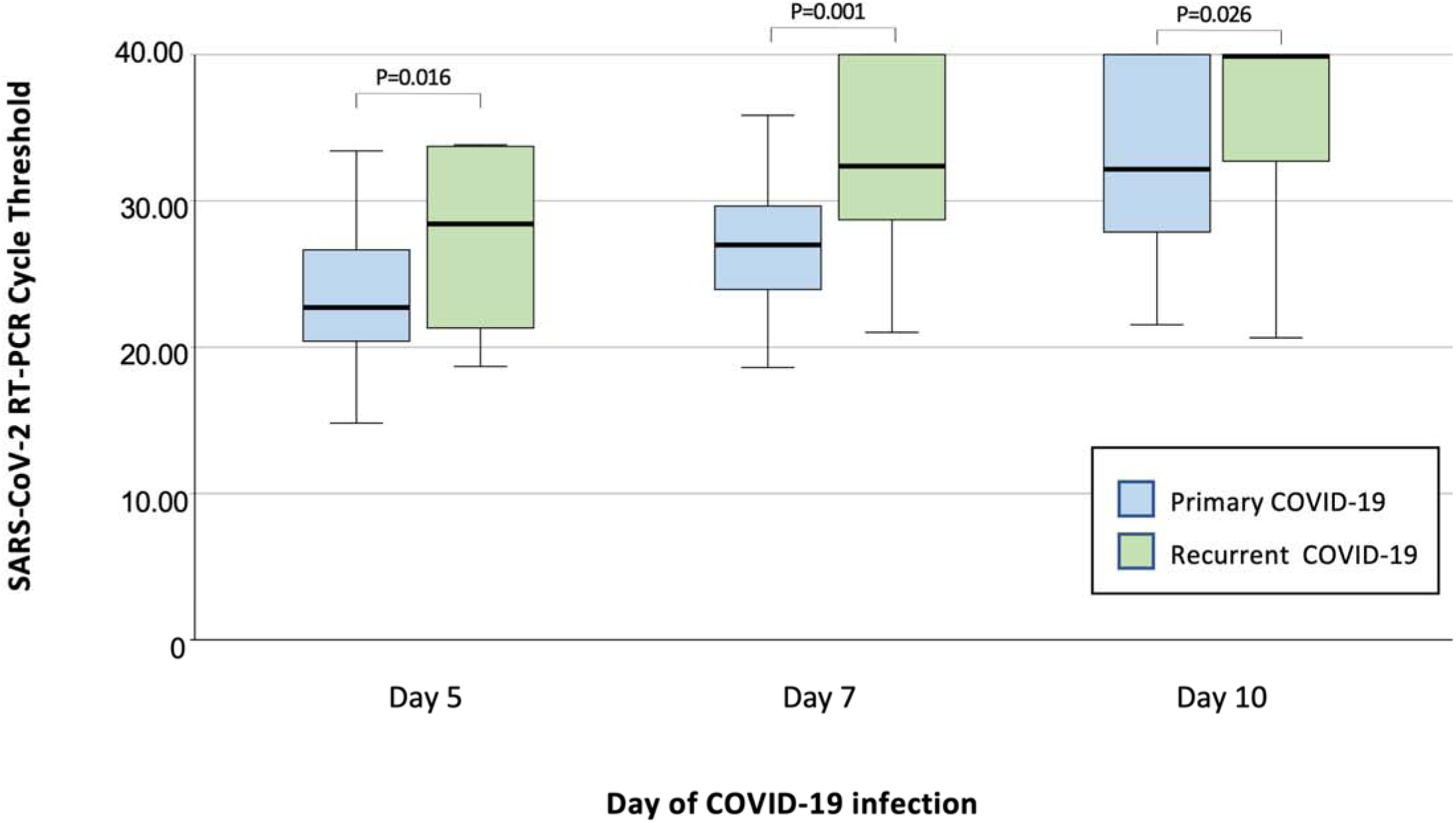
Box plot comparing *SARS-CoV-2 RT-PCR Cycle threshold* at day 5, 7 and 10 of primary vs. recurrent COVID-19 infection. Footnote: The horizontal line in each box indicates the median, whereas the top and bottom lines represent the 75th and 25th percentile, respectively. Error bars represent 95% confidence intervals. Negative RT-PCR results were attributed a Ct-value of 40 to facilitate data visualization. Comparison between primary vs recurrent infections assessed by Mann-Whitney U test.

**Table 3.** Comparison of rapid antigen detection test results of healthcare workers with primary vs recurrent COVID-19

|  | Day 5 of infection |  |  | Day 7 of infection |  |  | Day 10 of infection |  |  |
| --- | --- | --- | --- | --- | --- | --- | --- | --- | --- |
|  | Primary<br>COVID-19<br>N (%) | Recurrent<br>COVID-19<br>N (%) | P-value | Primary<br>COVID-19<br>N (%) | Recurrent<br>COVID-19<br>N (%) | P-value | Primary<br>COVID-19<br>N (%) | Recurrent<br>COVID-19<br>N (%) | P-value |
| **RADT result (n)** | 100 | 20 |  | 99 | 19 |  | 98 | 19 |  |
| Positive RADT | 86 (86.0) | 11 (57.9) | 0.005 | 73 (73.7) | 3 (15.8) | <0.001 | 40 (40.8) | 0 (0.0) | <0.001 |
| Negative RADT | 7 (7.0) | 6 (31.6) |  | 18 (18.2) | 16 (84.2) |  | 45 (45.9) | 19 (100) |  |
| Uncertain RADT | 7 (7.0) | 2 (10.5) |  | 8 (8.1) | 0 (0.0) |  | 13 (13.3) | 0 (0.0) |  |

Abbreviation: RADT, Rapid antigen detection test

In multivariate analysis (Table 4), the following variables were independently associated ongoing infectivity: A RT-PCR Ct value <23 was independently associated with an increased probability of infectivity on each follow-up visit (adjusted OR [aOR] on day 5, 22.75; p<0.001; aOR on day 7, 182.30; p<0.001; and aOR on day 10; 24.71; p=0.02). A Ct value ranging between 23 and 27 was also predictive of ongoing infectivity at Day 7 and Day 10. A history of previous COVID-19 was independently associated with a decreased probability of infectivity on day 5 (aOR, 0.005; p<0.001). By contrast, there was no significant association between ongoing infectivity and the absence of fever (with or without the use of antipyretics), symptom improvement, or RADT results.

**Table 4.** Predictors of infectivity among HCWs with COVID-19 (multivariate analysis)

|  | Day 5 (n=121) |  |  | Day 7 (n=117) |  |  | Day 10 (n=117) |  |  |
| --- | --- | --- | --- | --- | --- | --- | --- | --- | --- |
|  | Adjusted OR | 95% CI | P-value | Adjusted OR | 95% CI | P-value | Adjusted OR | 95% CI | P-value |
| **Female sex** | 0.42 | 0.09-2.06 | 0.287 | 1.28 | 0.31-5.34 | 0.73 | 0.83 | 0.16-4.18 | 0.82 |
| **Age (years)** |  |  |  |  |  |  |  |  |  |
| 20-39 | Ref |  |  | Ref |  |  | Ref |  |  |
| 40-59 | 0.50 | 0.15-1.68 | 0.26 | 1.43 | 0.47-4.34 | 0.52 | 1.28 | 0.36-4.63 | 0.70 |
| 60-77 | 0.17 | 0.02-1.63 | 0.12 | 0.52 | 0.06-4.71 | 0.56 | 2.54 | 0.25-26.31 | 0.43 |
| **Immunity status stratified by timing of last vaccine and previous COVID-19** |  |  |  |  |  |  |  |  |  |
| No previous infection & last vaccine dose ≥6 months ago | Ref |  |  | Ref |  |  | Ref |  |  |
| No previous infection & last vaccine dose <6 months ago | 0.27 | 0.03-2.33 | 0.23 | 7.50 | 0.89-62.83 | 0.06 | 1.41 | 0.14-14.15 | 0.77 |
| Previous infection, last vaccine dose > or < 6 months ago<sup>a</sup> | 0.005 | 0.002-0.16 | 0.003 | 0.14 | 0.003-6.61 | 0.32 | NE |  |  |
| **RADT result** |  |  |  |  |  |  |  |  |  |
| Negative | Ref |  |  | Ref |  |  | NE |  |  |
| Positive | 0.69 | 0.11-4.43 | 0.70 | 3.20 | 0.74-13.91 | 0.12 | NE |  |  |
| Uncertain | 0.14 | 0.1-1.48 | 0.10 | 0.07 | 0.002-1.82 | 0.11 | NE |  |  |
| **SARS-CoV-2 RT-PCR Ct** |  |  |  |  |  |  |  |  |  |
| ≥27 (including negative) | Ref |  |  | Ref |  |  | Ref |  |  |
| 23 - <27 | 1.30 | 0.29-5.62 | 0.73 | 4.81 | 1.52-15.25 | 0.008 | 12.39 | 3.32-46.20 | <0.001 |
| 14 - <23 | 22.75 | 3.89-133.05 | <0.001 | 182.30 | 8.83-3764.36 | 0.001 | 24.71 | 1.53-398.50 | 0.02 |
| **SARS-CoV-2 lineage<sup>b</sup>** |  |  |  |  |  |  |  |  |  |
| BA.1, BA.2 and subvariants | Ref |  |  | Ref |  |  | Ref |  |  |
| BA.4, BA.5, BQ.1, XBB and subvariants | 4.14 | 0.50-33.97 | 0.19 | 3.13 | 0.46-21.43 | 0.24 | 2.95 | 0.52-16.70 | 0.22 |
| Evolution of symptoms |  |  |  |  |  |  |  |  |  |
| Symptoms are better or entirely gone | Ref |  |  | Ref |  |  | NE |  |  |
| Symptoms are the same or worse than before | 0.52 | 0.11-2.57 | 0.42 | 18.67 | 0.98-355.49 | 0.05 | NE |  |  |
| Fever and antipyretic use<sup>c</sup> |  |  |  |  |  |  |  |  |  |
| No fever, without antipyretics use | Ref |  |  | Ref |  |  | Ref |  |  |
| No fever, with antipyretics use | 4.83 | 1.30-17.98 | 0.85 | 1.32 | 0.40-4.35 | 0.65 | 4.16 | 1.00-16.95 | 0.047 |
| Fever | 1.21 | 0.18-8.17 | 0.85 | NA |  |  | NA |  |  |

<sup>a</sup> Regardless of timing of last vaccine dose
<sup>b</sup> For 9 individuals with missing information, lineage BA.1 / BA.2 or lineage BA.4 / BA.5 / BQ.1 / XBB were assigned based on circulating variants at the date of testing
<sup>c</sup> For the analyses of day 7 and day 10, “fever” and “no fever, with antipyretic use” were considered a single category
Abbreviations: CI, confidence interval; NE, not estimable; NA, not applicable; OR, odds ratio; RADT, Rapid antigen detection test; Ref, reference category; RT-PCR Ct, cycle threshold of real-time polymerase chain reaction

### Performance of return-to-work algorithms

We applied the US CDC criteria to our cohort to identify non-infectious individuals on day 7 of COVID-19 (Figure 4).^7^ Approximately three quarters (88/117; 75.2%) would be ineligible for return to work because of fever (n=3), the use of antipyretics (n=35), a lack of symptom improvement (n=3) or a positive RADT (n=47). Hence, only 29 HCWs (24.8%) would meet all the return-to-work criteria. Of these, 17.2% (5/29) were infectious by viral culture, and 82.8% (24/29) were non-infectious. Hence, this algorithm could identify a third (35.8%; 24/67) of all non-infectious individuals on day 7. If all 29 HCWs who fulfilled all criteria returned to work on day 7, this algorithm would decrease absenteeism by 7.4%.

**FIGURE 4.**
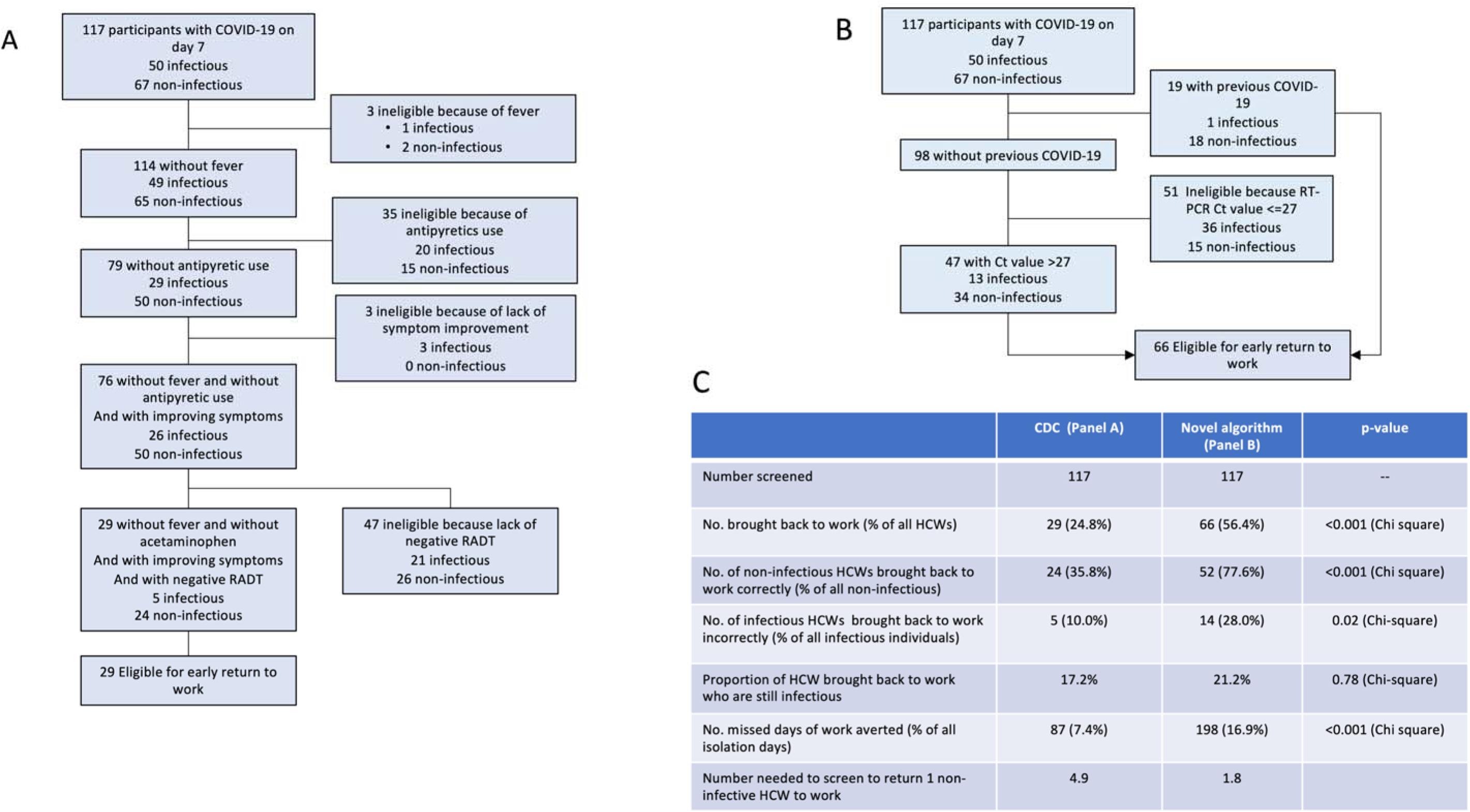
Performance of return-to-work criteria for healthcare workers with COVID-19. Panel A shows the performance of the US Centers for Diseases Control and Prevention (US CDC) Return to Work criteria on a cohort of healthcare workers with COVID-19. Panel B shows the performance of an alternative set of criteria derived from the current study. Panel C compares the CDC and alternative criteria.

We applied an alternative algorithm that used a history of previous COVID-19 and a RT-PCR Ct value >27 to predict loss of infectivity on day 7. This algorithm would identify 56.4% (66/117) of all HCWs as eligible for return to work and could avoid 198 days of absence (16.9%). Of these, 52 (78.8%) were non-infectious, and 14 (21.2%) were infectious. This algorithm would identify a greater proportion of all non-infectious HCWs than the CDC algorithm (77.6% vs 35.8%; p<0.001). Even though it would return to work a greater absolute number of infectious individuals, it would not significantly increase in the probability of returning to work an infectious HCW (21.2% vs 17.2% of all eligible HCWs, p=0.78).

Given that approximately two thirds of individuals with recurrent COVID-19 were non-infectious by day 5, we explored various criteria that could accelerate their return-to-work (eFigure 1 in supplementary appendix). Among these, a RT-PCR Ct value >27 could identify most (77%; 10/13) non-infectious individuals on day 5 with low probability (9%; 1/11) of returning to work an infectious HCW.

## Discussion

Healthcare workers with COVID-19 must be removed from work, but their absence can exacerbate staff shortages.^22^ There is a need to identify predictors of loss of infectivity of SARS-CoV-2 to prevent unwarranted prolongation of absence. Most published studies on this topic have relatively low number of participants limiting the identification of predictors of loss of infectivity.^6, 23, 24^

In this prospective study, approximately three quarters (71.9%) were still shedding infectious viral particles on 5^th^ day of infection, half (46.7%) on the 7^th^ day, and a fifth (18.2%) on the 10^th^ day. These results, along with other recent publications,^25–28^ differ from those of earlier studies that estimated the duration of infectivity to 10 days or less.^6, 29^ A study of 66 individuals infected with the Omicron variant BA.1 reported that a quarter of participants were still infective on the 10^th^ day of infection.^9^ In another recent study, 8.5% were still shedding viable virus on day 14.^30^

However, our study also identifies important nuances regarding durations of infectivity. Nowadays, an increasing proportion of infections occurs in individuals who have hybrid immunity due to vaccination and previous COVID-19. Our study identified that vaccinated individuals with recurrent COVID-19 have a significantly shorter duration of infectivity as well as a distinct viral kinetic as evidenced by lower viral loads and earlier negativisation of RADT. A prepublication study of 1400 professional athletes also reported faster viral clearance by RT-PCR in individuals with recurrent COVID-19 compared with primary infections (4.9 vs 7.2 days, respectively).^31^ A cohort study in Alaska recently reported that individuals with previous COVID-19 infections were less likely to have a positive RADT result by day 5 compared to those with primary COVID-19.^25^ A similar phenomenon has be described with other Coronaviridae. A study performed three decades ago among volunteers infected with coronavirus 229E determined that reinfections led to a shorter duration of virus shedding compared with the primary episode.^32^ Taken together, these findings profoundly alter our understanding of the infectivity and viral kinetic of recurrent COVID-19.

Our study also identified that a higher RT-PCR Ct value was the strongest independent predictor of loss of infectivity. This confirms findings from other studies that demonstrated that higher Ct values correspond with a non-replicative virus^3, 26, 33–39^. Hence, RT-PCR Ct values could help determine the timing of return to work of HCWs. A negative RADT result was also predictive of loss of infectivity by bivariate analysis.^40^ However, our multivariate analysis indicates that RT-PCR Ct values hold superior predictive capacity.

Our study also suggests that the guidance provided by the CDC to accelerate the return to work of infected HCWs is relatively stringent as it allows the return to work of only a third of all non-infectious individuals.^7, 8^ Also, they appear to have moderate discriminatory power as up to a sixth of those eligible for return to work are still shedding viable virus. By contrast, an alternative algorithm using RT-PCR Ct values and a history of previous COVID-19 could be able to return to work a greater proportion of HCWs on Day 7 without significantly increasing the probability of returning to work an infectious HCW. Importantly, the performance of these algorithms will be influenced by whether the COVID-19 episode is a first episode or a recurrence. In our opinion, such distinction will be essential when updating these algorithms.

Our study has strengths. To our knowledge, this study is the first to demonstrate that individuals with recurrent COVID-19 have significantly shorter durations of infectivity using the gold standard of viral culture. It is among the largest that assessed infectivity using viral culture. Our laboratory technique was sensitive and could detect viable viruses in many individuals 10 days into their infection. It also has limitations. The study enrolled young, healthy, and highly immunized participants with mild COVID-19, so generalizability to other populations is uncertain. Even though culture positivity is the best marker of infectivity, the correlation between culture positivity and transmission remains unclear.^9^ Additional studies, including validation with an external cohort, would be required to better inform return-to-work policies.^20, 21^

In conclusion, our study detected a higher RT-PCR Ct value and COVID-19 reinfection as independent predictors of loss of infectivity in a highly vaccinated population, and suggests that return-to-work algorithms could be optimized to limit absenteeism.^10^ Further studies are needed to further characterize the viral kinetics of COVID-19 reinfections as they appear to fundamentally differ from those with primary COVID-19.

## Data Availability

All data produced in the present work are contained in the manuscript

## Acknowledgements

Conflict of Interest Disclosures: Dr Longtin reported receiving a grant from Summit (Oxford) outside the submitted work. No other disclosures were reported.

## Funding/Support

This work was supported by the Ministère de la Santé et des Services Sociaux du Québec. The sponsor was not involved in the design and conduct of the study; collection, management, analysis, and interpretation of the data; preparation, review, or approval of the manuscript; and decision to submit the manuscript for publication.

## Data Access, Responsibility, and Analysis

Yves Longtin had full access to all the data in the study and takes responsibility for the integrity of the data and the accuracy of the data analysis. Data not publicly available.

## Author Contribution

Concept and Design: Dzieciolowska, Fafard, Charest, De Serres, J. Longtin, Villeneuve, Y. Longtin Acquisition, analysis, or interpretation of data: Dzieciolowska, Roy, Charest, Fafard, Levade, Beaulac, Parkes, Y. Longtin

Drafting of manuscript: Dzieciolowska, Y. Longtin

Critical revision of the manuscript for important intellectual content: Charest, Roy, Fafard, Carazo, Levade, J. Longtin, Parkes, Beaulac, Villeneuve, Savard, Corbeil, De Serres,

Statistical analyses: Carazo, De Serres, Y. Longtin

Obtained funding: Y. Longtin, De Serres

Administrative, technical or material support: Dzieciolowska, Y. Longtin

Supervision: Y. Longtin, De Serres

## Notes

### Competing Interest Statement

Y. Longtin reports receiving research support from Summit (Oxford). All other co-authors report no potential conflicts of interest.

### Funding Statement

This work was funded by the Ministere de la Sante et des Services Sociaux du Quebec. The funding source had no role in the design of the study, execution, analyses, interpretation of data, writing of report or decision to submit the manuscript.

### Author Declarations

Ethics committee/IRB of the Jewish General Hospital Sir Mortimer B Davis gave ethical approval for this work

