## Appendix for "Timing and Predictors of Loss of Infectivity among Healthcare Workers with Primary and Recurrent COVID-19: a Prospective Observational Cohort Study"

**eMethods.** Whole Genome Sequencing

**eFIGURE 1.** Performance of return-to-work criteria for healthcare workers with recurrent COVID-19 on the fifth day of their infection

This supplemental material has been provided by the authors to give readers additional information about their work.
**eMethods**

**Whole Genome Sequencing and Sequence Data Analysis**

Nucleic acids were extracted from 0.2-mL aliquots using the NUCLISENS easyMAG (bioMérieux, Marcy-lʼÉtoile, France). The viral RNA was sequenced using Illumina technology, and a targeted SARS-CoV-2 amplification strategy was employed based on the ARTIC V4.1 primer scheme.^1^ Libraries were prepared with the Illumina COVIDSeq Test kit according to manufacturer’s recommendation^2^ and sequenced with a NextSeq 1000 (Illumina). Data analysis was conducted using the GenPipes Covseq pipeline,^3^ which performed alignment and produced variant calls. Initially, host reads were removed by aligning them to a hybrid reference consisting of human (GRCh38) and Wuhan-Hu-1 SARS-CoV-2 reference (MN908947.3) sequences. Raw reads were trimmed using cutadapt (v2.10), and then aligned to the reference using bwa-mem (v0.7.17).^4^ The resulting aligned reads were filtered using sambamba (v0.7.0),^5^ which removed paired reads with insert sizes outside the 60-300 bp range, unmapped reads, and all secondary alignments. Remaining ARTIC primers (v4.1) were trimmed using iVar (v1.3).^6^ To generate a consensus sequence, a pileup was produced using Samtools (v1.12),^7^ which was then used as input for FreeBayes (v1.3.4) to create a consensus sequence for regions with a minimum of 10× depth and using reads with a Q score > 20.^8^ Mutations were annotated with snpEff (v4.5).^9^ Single nucleotide variants below 5% allele frequency were filtered out. A full description of the process can be found here:

<https://c3g.github.io/covseq_McGill/SARS_CoV2_Sequencing/Illumina_overview.html>.

**Variant identification and detection of recombination**

Variant identification was performed using the Pangolin program (v4.2, UShER analysis mode),^10^ and the program ncov-recombinant (v.0.6.0) was used to characterize recombinant lineages non-identified by Pangolin.^11^

**eMethods References**

8.	Garrison E, Marth G. Haplotype-based variant detection from short-read sequencing. *arXiv preprint arXiv:12073907*. 2012;

9.	Cingolani P, Platts A, Wang le L, et al. A program for annotating and predicting the effects of single nucleotide polymorphisms, SnpEff: SNPs in the genome of Drosophila melanogaster strain w1118; iso-2; iso-3. *Fly (Austin)*. Apr-Jun 2012;6(2):80-92. doi:10.4161/fly.19695

10.	O'Toole A, Scher E, Underwood A, et al. Assignment of epidemiological lineages in an emerging pandemic using the pangolin tool. *Virus Evol*. 2021;7(2):veab064. doi:10.1093/ve/veab064

11.	ktmeaton/ncov-recombinant. <https://github.com/ktmeaton/ncov-recombinant>, 2023.

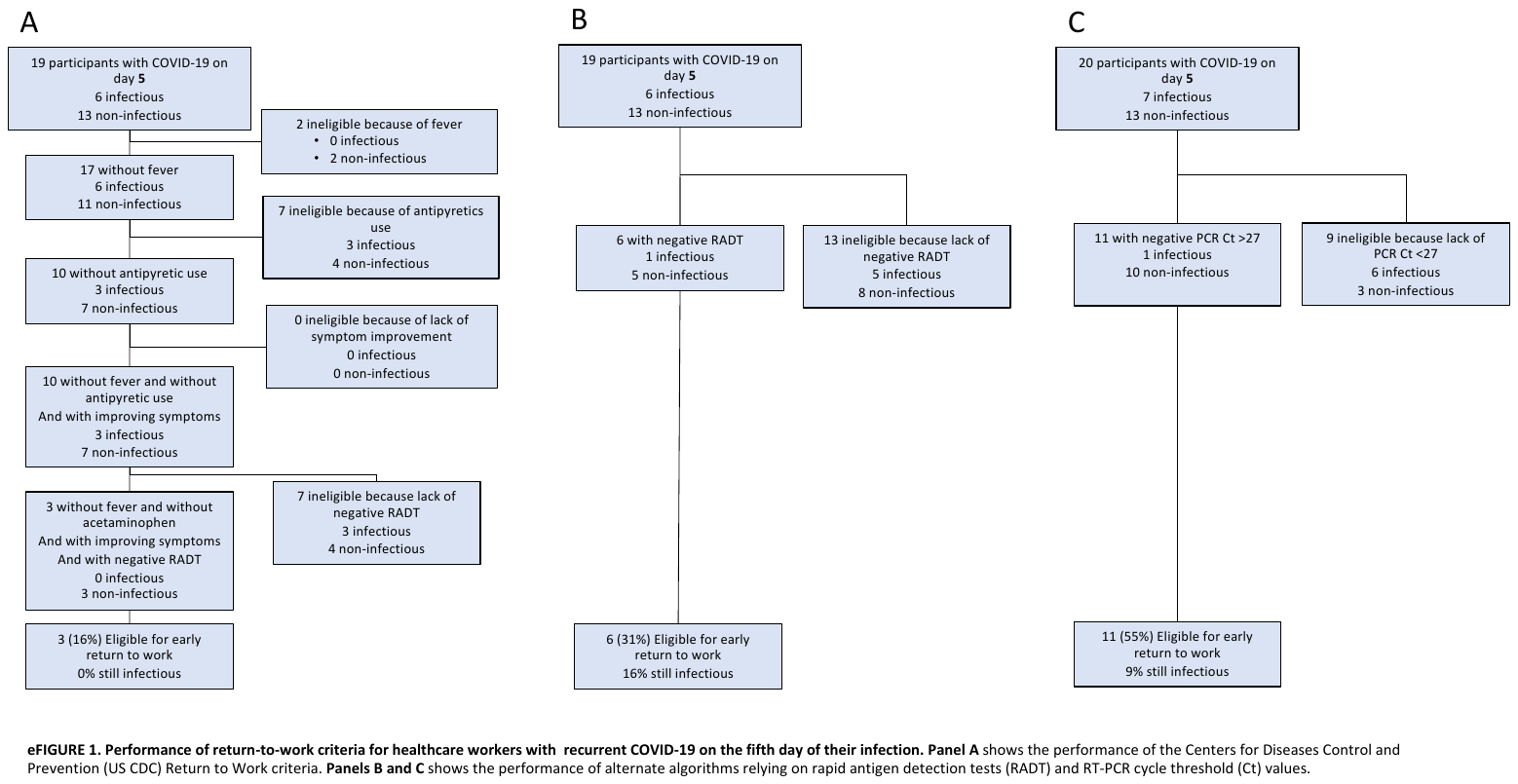
